# Cumulative incidence and diagnosis of SARS-CoV-2 infection in New York

**DOI:** 10.1101/2020.05.25.20113050

**Authors:** Eli S. Rosenberg, James M. Tesoriero, Elizabeth M. Rosenthal, Rakkoo Chung, Meredith A. Barranco, Linda M. Styer, Monica M. Parker, Shu-Yin John Leung, Johanne E. Morne, Danielle Greene, David R. Holtgrave, Dina Hoefer, Jessica Kumar, Tomoko Udo, Brad Hutton, Howard A. Zucker

## Abstract

**Importance:** New York State (NYS) is an epicenter of the United States’ COVID-19 epidemic. Reliable estimates of cumulative incidence of SARS-CoV-2 infection in the population are critical to tracking the extent of transmission and informing policies, but US data are lacking, in part because societal closure complicates study conduct.

**Objective:** To estimate the cumulative incidence of SARS-CoV-2 infection and percent of infections diagnosed in New York State, overall and by region, age, sex, and race and ethnicity.

**Design:** Statewide cross-sectional seroprevalence study, conducted April 19-28, 2020.

**Setting:** Grocery stores (n=99) located in 26 counties throughout NYS, which were essential businesses that remained open during a period of societal closure and attract a heterogenous clientele.

**Participants:** Convenience sample of patrons ≥18 years and residing in New York State, recruited consecutively upon entering stores and via an in-store flyer.

**Exposures:** Region (New York City, Westchester/Rockland, Long Island, Rest of New York State), age, sex, race and ethnicity.

**Main Outcomes:** Primary outcome: cumulative incidence of SARS-CoV-2 infection, based on dry-blood spot (DBS) SARS-CoV-2 antibody reactivity; secondary outcome: percent of infections diagnosed.

**Results:** Among 15,101 adults with suitable DBS specimens, 1,887 (12.5%) were reactive using a validated SARS-CoV-2 IgG microsphere immunoassay (sensitivity 87.9%, specificity 99.75%). Following post-stratification weighting on region, sex, age, and race and ethnicity and adjustment for assay characteristics, estimated cumulative incidence through March 29 was 14.0% (95% CI: 13.3-14.7%), corresponding to 2,139,300 (95% CI: 2,035,800-2,242,800) infection-experienced adults. Cumulative incidence was higher among Hispanic/Latino (29.2%, 95% CI: 27.2-31.2%), non-Hispanic black/African American (20.2% 95% CI, 18.1-22.3%), and non-Hispanic Asian (12.4%, 95% CI: 9.4-15.4%) adults than non-Hispanic white adults (8.1%, 95% CI: 7.4-8.7%, p<.0001). Cumulative incidence was highest in New York City (NYC) 22.7% (95% CI: 21.5%-24.0). Dividing diagnoses reported to NYS by estimated infection-experienced adults, an estimated 8.9% (95% CI: 8.4-9.3%) of infections were diagnosed, with those ≥55 years most likely to be diagnosed (11.3%, 95% CI: 10.4-12.2%).

**Conclusions and Relevance:** Over 2 million adults were infected through late March 2020, with substantial variations by subpopulations. As this remains below herd immunity thresholds, monitoring, testing, and contact tracing remain essential public health strategies.

## Introduction

The first cases of COVID-19 were identified in New York State (NYS) in early March, 2020 and since then NYS, particularly the metropolitan New York City (NYC) area, has become one of the most-impacted communities in the United States.^1,2^ As of May 9, 2020, over 335,000 laboratory-confirmed diagnoses have been made, accounting for approximately 25% of US diagnoses.^2,3^ As with most infections, lab-confirmed diagnoses undercount the true population-level burden of infections; with SARS-CoV-2, the virus that causes COVID-19, key factors that contribute to underdiagnosis include absent or mild symptoms and access to testing.^4^ Thus although NYS has tested more residents for COVID-19 than any other state (over 1,180,000 persons tested through May 9, 2020), it is likely that laboratory-confirmed cases represent a relatively small portion of the total number of persons with a history of infection in NYS.^3^

Estimates of COVID-19 cumulative incidence (i.e. prevalence of previous or current infection) can inform the extent of epidemic spread as well as the number of persons still susceptible and progress towards herd immunity, which are critical for parameterizing simulation models and informing policies, including those for altering societal restrictions.^5^ Furthermore, such data provide needed denominators for understanding the extent of diagnosis and rates of hospitalization, morbidity, and mortality, and geographic differences.

Antibody testing for SARS-CoV-2 has emerged as an important tool for understanding infection history. Although a several-week window period for development of IgG antibodies and evidence that not all persons with infection develop an antibody response limits their utility for diagnostics, and their interpretation for short- and long-term immunity remain uncertain, as with other infections, antibody prevalence serostudies with validated assays can assess population-level cumulative incidence in the recent past.^6-10^

Antibody serostudies for SARS-CoV-2 are being conducted in other countries and in the US are occurring on the national and county levels, but none have been conducted at the state level and no findings have been peer-reviewed.^11,12^ The current array of recommendations against individual movement and business operation during the pandemic complicates study specimen collection. A recent RNA survey in Iceland and serosurvey in Santa Clara County, California, conducted sampling at centralized testing sites, which offer ease of execution particularly in small geographies, with potentially large self-selection biases.^11,13^ Alternative approaches include random at-home mail-in testing and community-intercept studies in high-traffic locations that remain open.^12^

To provide a statewide picture of COVID-19 infection through late-March and diagnoses by early-April 2020, during April 19-28 2020, the NYS Department of Health (NYSDOH) conducted a community-based serostudy throughout NYS. Cumulative incidence by geographic and demographic features was estimated from weighted reactivity rates that were adjusted for validated test characteristics. Combining these findings with cumulative diagnoses enabled estimation of the percent of infections diagnosed.

## Methods

### Field study

The NYS DOH conducted a convenience sample of over 15,000 New Yorkers attending 99 grocery stores across 26 counties, which contain 87.3% of the state’s population, located in all regions of NYS (Figure). Grocery stores were chosen as the testing venue because they were classified an essential business to remain open and, due to the necessity of grocery shopping, they attract a heterogeneous clientele.^14^ Store locations were chosen to increase sample coverage of the racial and ethnic diversity of the statewide population.

**Figure.**
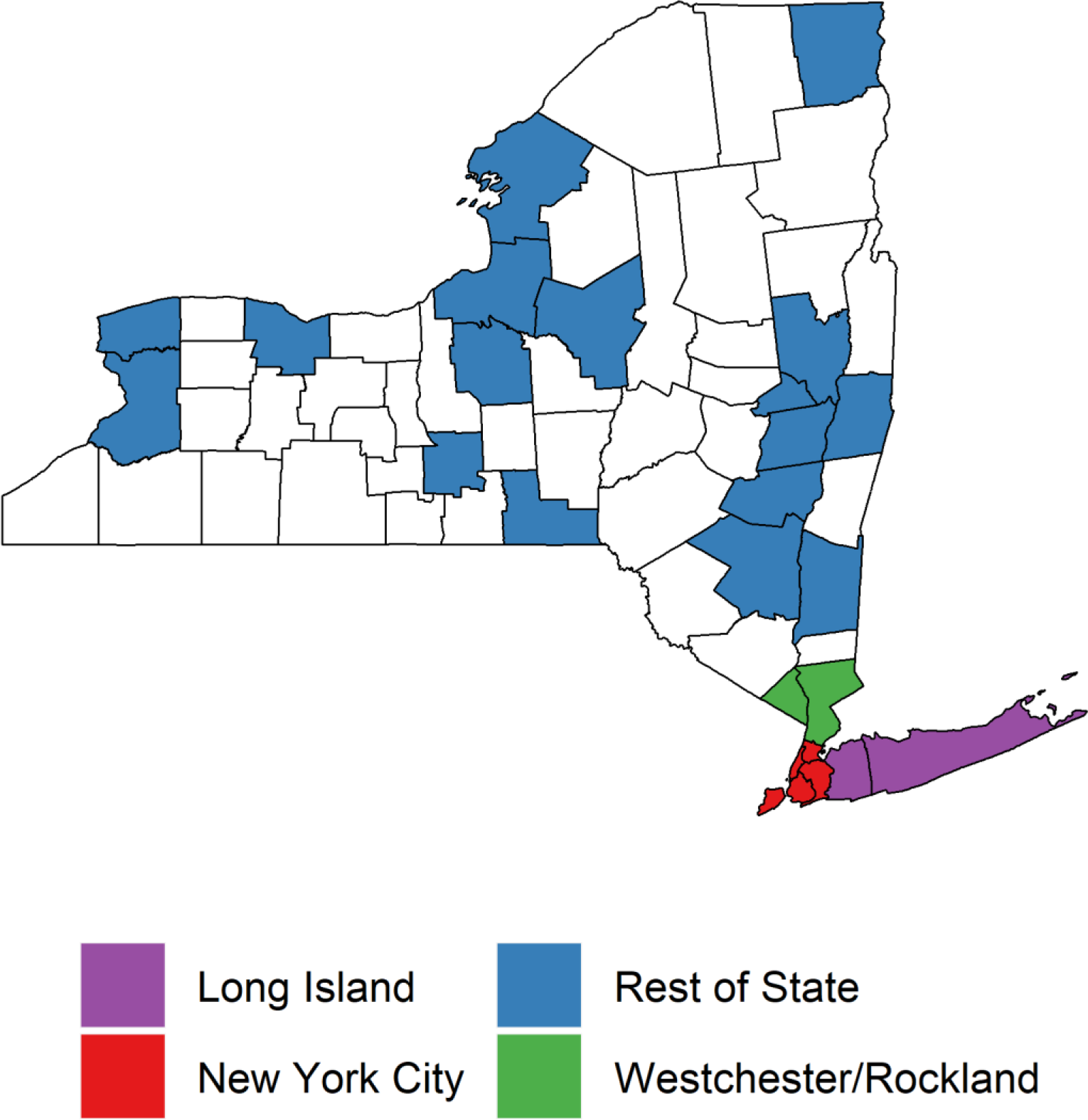
New York State counties included in the New York State Department of Health Serological Testing Survey^1^

Testing occurred over 6 distinct days from 4/19/2020 through 4/28/2020. Each store had a team of 6-8 staff responsible for recruiting participants, collecting specimens, recording data, and managing specimen transport to Wadsworth Center Laboratory (Albany, NY) for analysis. Eligible subjects were adults ≥18 years, New York residents irrespective of county, recruited through a recruitment flyer posted at stores and by systematically approaching each patron as they entered the store. Patrons were given information about the testing and if interested, completed written informed consent. Procedures included a brief demographic questionnaire and dried blood spot (DBS) collection by trained personnel. Approximately 13% of participants initially had missing demographic data. Staff attempted to capture these data through >2,500 follow-up phone calls, reaching all but approximately 75 participants, who were subsequently excluded from analyses. Test results were delivered to participants by text-message if non-reactive and by phone if indeterminant or reactive.

### Testing approach

Blood was collected by fingerstick onto custom 903 filter paper cards labeled with a specimen ID. Cards were dried for 3-4 hours at ambient temperature and transported to the Wadsworth Center. A fully saturated ≥3-mm diameter DBS was required. A total of 525 DBS cards from eligible individuals were rejected; 433 with insufficient or improperly collected blood, 92 with no specimen ID. Acceptable DBS cards were processed for testing.

SARS-CoV IgG testing was conducted using a microsphere immunoassay (MIA) developed and validated for DBS by the NYSDOH Wadsworth Center. Briefly, nucleocapsid (N) antigen-coupled magnetic beads were incubated with blood eluted from a 3-mm DBS punch. Phycoerythrin-labeled goat anti-human IgG secondary antibody was used to detect microsphere-bound IgG antibodies and median fluorescence intensity (MFI) was determined using a FlexMap 3D (Luminex Corp., Austin, TX). The mean MFI of 90-100 negative DBS was used to set cut-offs; results greater than the mean MFI plus 6 standard deviations (SD) were reported as reactive; results less than the mean MFI + 3 SD were nonreactive and results between mean MFI +3 to +6 were indeterminate. Serosurvey testing was initiated with SARS-CoV IgG v1 which used SARS-CoV-1 N antigen (Wadsworth Center, Albany, NY), and was completed using SARS-CoV IgG v2 which used SARS CoV-2 N protein (Sino Biological, Wayne, PA) after validation studies confirmed comparable performance.

Assay validation studies are described in eTables 1-5 and eFigure in the Supplement. Based on testing DBS collected prior to December 2019, specificity was estimated between 99.5% (95% CI: 98.5-100%) to 100% (95% CI: 96.1-100%). Serum collected from individuals diagnosed with non-COVID-19 respiratory and non-respiratory agents were tested to assess cross-reactivity; only 1 of 85 samples was reactive. Of 232 SARS CoV-2 PCR-positive DBS collected a median 35 days post-symptom onset, 204 (87.9%, 95% CI: [83.7-92.1%]) were reactive, informing sensitivity and thus incorporating both test performance and the proportion of infected persons who never develop IgG.^6,7^

## Analysis

We estimated SARS-Cov-2 cumulative incidence from observed antibody reactivity using two sequential steps: 1) post-stratification weighting to standardize to the New York State population and 2) adjustment by estimated antibody test characteristics.

Using the National Center for Health Statistics bridged-race file, weights were assigned to each participant based on their membership in each of 160 strata of sex, race and ethnicity (Hispanic, non-Hispanic white, non-Hispanic black, non-Hispanic Asian, and non-Hispanic other), age (18-34, 35-44, 45-54, ≥55 years), and residential region (New York City, Westchester/Rockland, Long Island, Rest of State [ROS]).^15^ Post-stratification weights were defined as the proportion each stratum is represented in the state’s population divided by the analogous proportion in the sample.^16,17^ Next, we computed weighted frequencies for the percent reactive statewide, with one-way stratifications by sex, race and ethnicity, age group, and region, and two-way stratifications within levels of region, including 95% confidence intervals (CI), with differences assessed using Rao-Scott χ^2^ tests.^18^ Indeterminate results were assumed non-reactive and statistical procedures were two-sided at α=0.05.

In the second step, weighted reactivity estimates *(p_reactive_)* and their 95% CI bounds were corrected for test sensitivity and specificity, based on validation data, to yield cumulative incidence, per Bayes’ Rule as applied to the diagnostic 2×2 table: 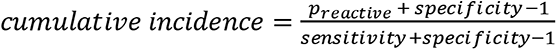.^11,19^ Primary analyses used the sensitivity and specificity point-estimates from the validation studies, with sensitivity analyses at the extremes of test characteristics’ 95% CI ([96.1% specificity, 92.1% sensitivity], [100% specificity, 83.7% sensitivity]). Test-characteristic adjusted cumulative incidence values were multiplied by the one- and two-way non-institutionalized adult populations from the American Community Survey 2014-2018 Public Use Microdata Sample file.^20^ This yielded the estimated total ‘infection-experienced’ adults with SARS-CoV-2 within each stratum, accounting for excluded settings such as prisons and nursing homes. With a study mid-point of April 23, and literature estimates of mean 4 days from infection to symptom onset and mean 21 days from onset to IgG detection, results represent cumulative incidence through approximately March 29.^6,8,21^

In NYS, diagnostic testing for SARS-CoV-2 is mandatorily reported electronically to NYSDOH. Using cumulative diagnoses reported and total numbers of infection-experienced adults, we estimated the percent of infections diagnosed overall and by region, sex, and age. For primary analyses, we accumulated diagnoses through April 9, based on the March 29 final infection date, 4 days to symptom onset, and mean 7 days from onset to diagnosis. Supplemental upper-bound estimates used the last plausible diagnosis date of May 8^th^, based on the April 28 final study day, 4 days being earliest time from onset to IgG detection and allowing PCR detection up to 14 days post-onset.^8^

## Results

Across NYS, a total of 15,626 adult residents with complete data were tested, of whom 15,101 (96.6%) had suitable specimens, of which 1,887 (12.5%) were reactive and 340 (2.3%) indeterminate. Following weighting, 12.5% were estimated reactive and following further adjustment for test characteristics, estimated cumulative incidence was 14.0% (95% CI: 13.3-14.7%), corresponding to 2,139,300 (95% CI: 2,035,800-2,242,800) infection-experienced adults in NYS through approximately March 29 (Table 1). In sensitivity analyses at the extremes of test characteristics, cumulative incidence ranged from 9.8% (95% CI: [9.1-10.5%]) to 15.0% (95% CI: [14.3-15.7%]), representing a total of 1,494,700 (95% CI: [1,391,800-1,597600]) to 2,286,600 (95% CI: [2,178,200-2,395,100]) adults in NYS (eTables 6-7).

**Table 1.**
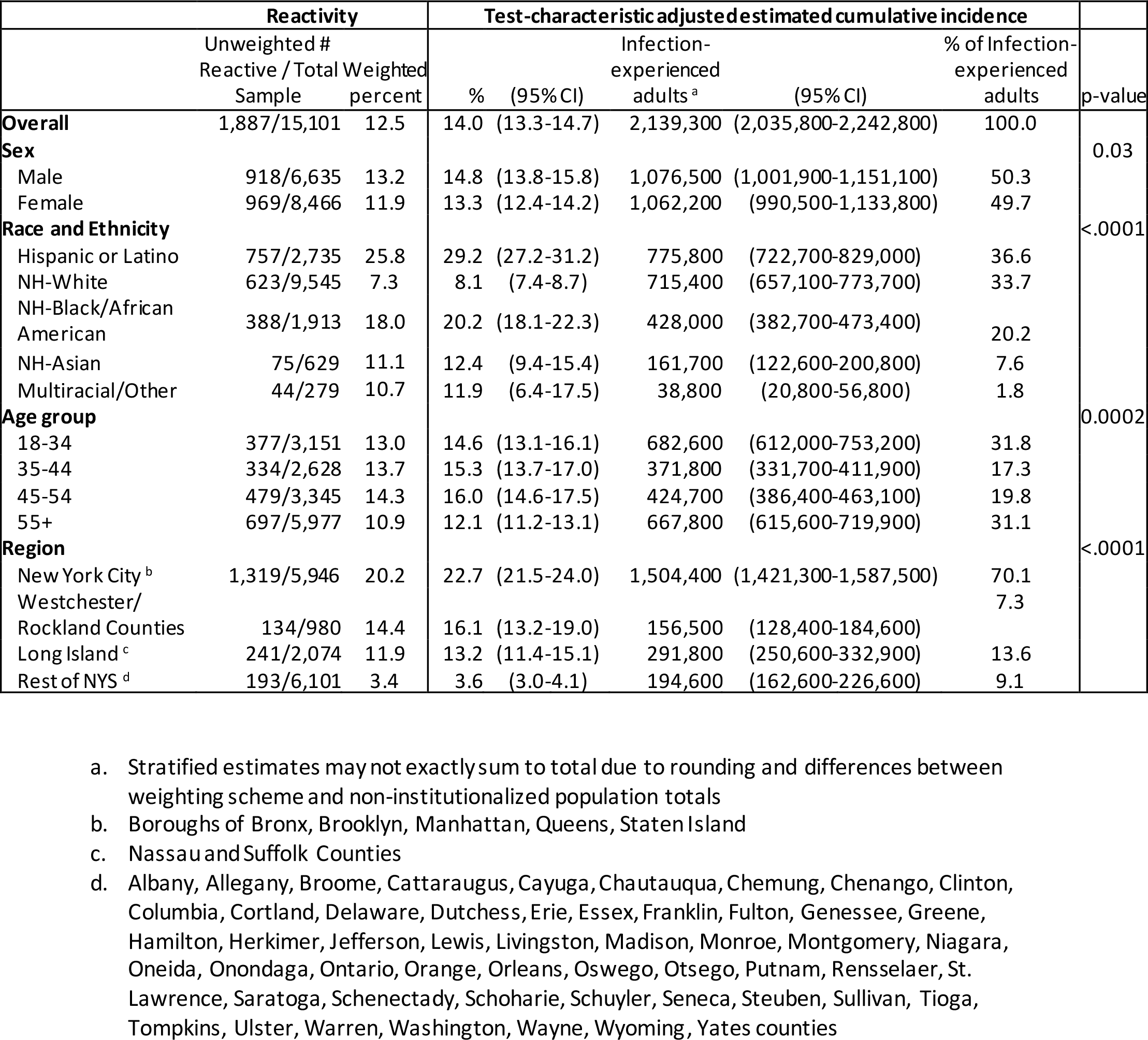
Reactivity and test-characteristic adjusted cumulative incidence of COVID-19, overall and by demographic factors and region

Cumulative incidence was higher among males (14.8%, 95% CI: [13.8-15.8%]) than females (13.3%, 95% CI: [12.4-14.2%], p=0.03), with males comprising 50.3% of adult infections. This differed significantly by race and ethnicity, with Hispanic/Latino (29.2%, 95% CI: 27.2-31.2%), non-Hispanic black/African American (20.2% [95% CI, 18.1-22.3%]), and non-Hispanic Asian (12.4%, 95% CI: [9.4-15.4%]) adults having higher cumulative incidence than non-Hispanic white adults (8.1%, 95% CI: [7.4-8.7%], p<.0001). Given these differences, Hispanics comprised the plurality (36.6%) of infection-experienced adults. Significant differences were also observed by age (p=0.0002), ranging from highest levels among persons 45-54 years old (16.0%, 95% CI: [14.6-17.5%]) to lowest among persons ≥55 years (12.1% [95% CI: 11.2-13.1%]).

We observed regional heterogeneity in cumulative incidence, ranging from 22.7% (95% CI: 21.5%-24.0%) in NYC residents, to 16.1% (95% CI: 13.2-19.0%) and 13.2% (11.4-15.1%) in the respective metropolitan areas of Westchester/Rockland Counties and Long Island, to 3.6% (95% CI: [3.0-4.1]) in ROS (p<0.0001). Demographic patterns were heterogenous by region (Table 2). Males had significantly higher cumulative incidence in all regions outside of, but not within NYC. The patterns of racial disparity observed statewide were similar and statistically significant within NYC, Westchester/Rockland, and Long Island, but not in ROS. In each of the former 3 regions, Hispanic/Latino persons represented >37% of infection-experienced adults, whereas in the latter non-Hispanic whites comprised a majority of infection-experienced adults (79.4%).

**Table 2.**
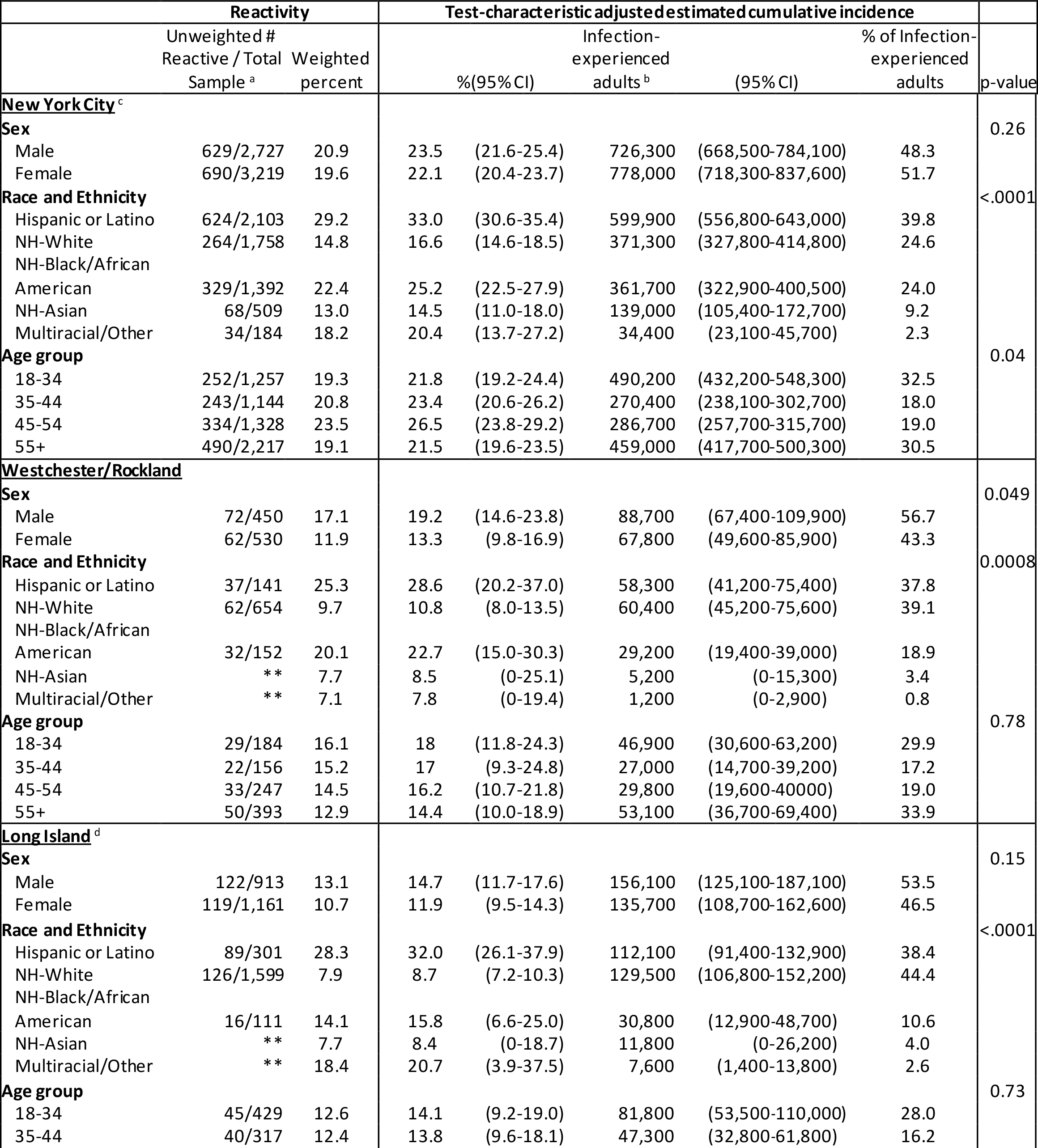
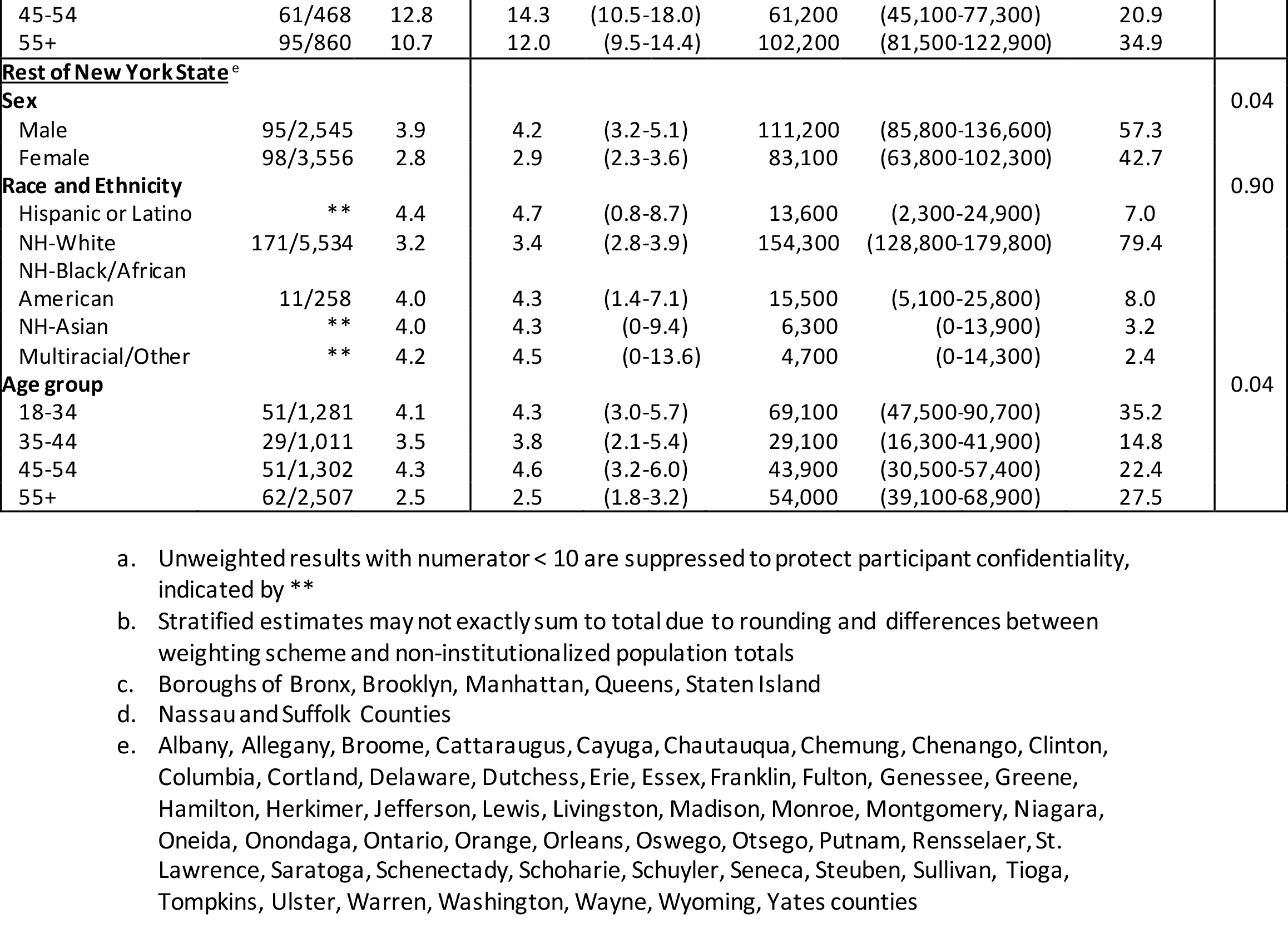
Reactivity and test-characteristic adjusted cumulative incidence of COVID-19, demographic factors within region

An estimated 8.9% (95% CI: 8.4-9.3%) of infections in NYS were diagnosed as of April 9^th^ 2020 (Table 3). Males (9.4%, 95% CI: 8.8-10.1%) had higher diagnosis levels than females (8.2%, 95% CI: 7.7-8.8%)). Those ≥55 years were most likely to be diagnosed (11.3%, 95% CI: 10.4-12.2%). Diagnosis rates in NYC (7.1%, 95% CI: 6.7-7.5%) and ROS (7.5%, 95% CI: 6.4-8.9%) were about half those observed in the other regions. Considering the May 8 upper-bound for diagnoses, a maximum of 15.7% (95% CI: 15.0-16.5%) of overall infections could have been diagnosed, with similar patterns observed across levels of each factor (eTable 8).

**Table 3.**
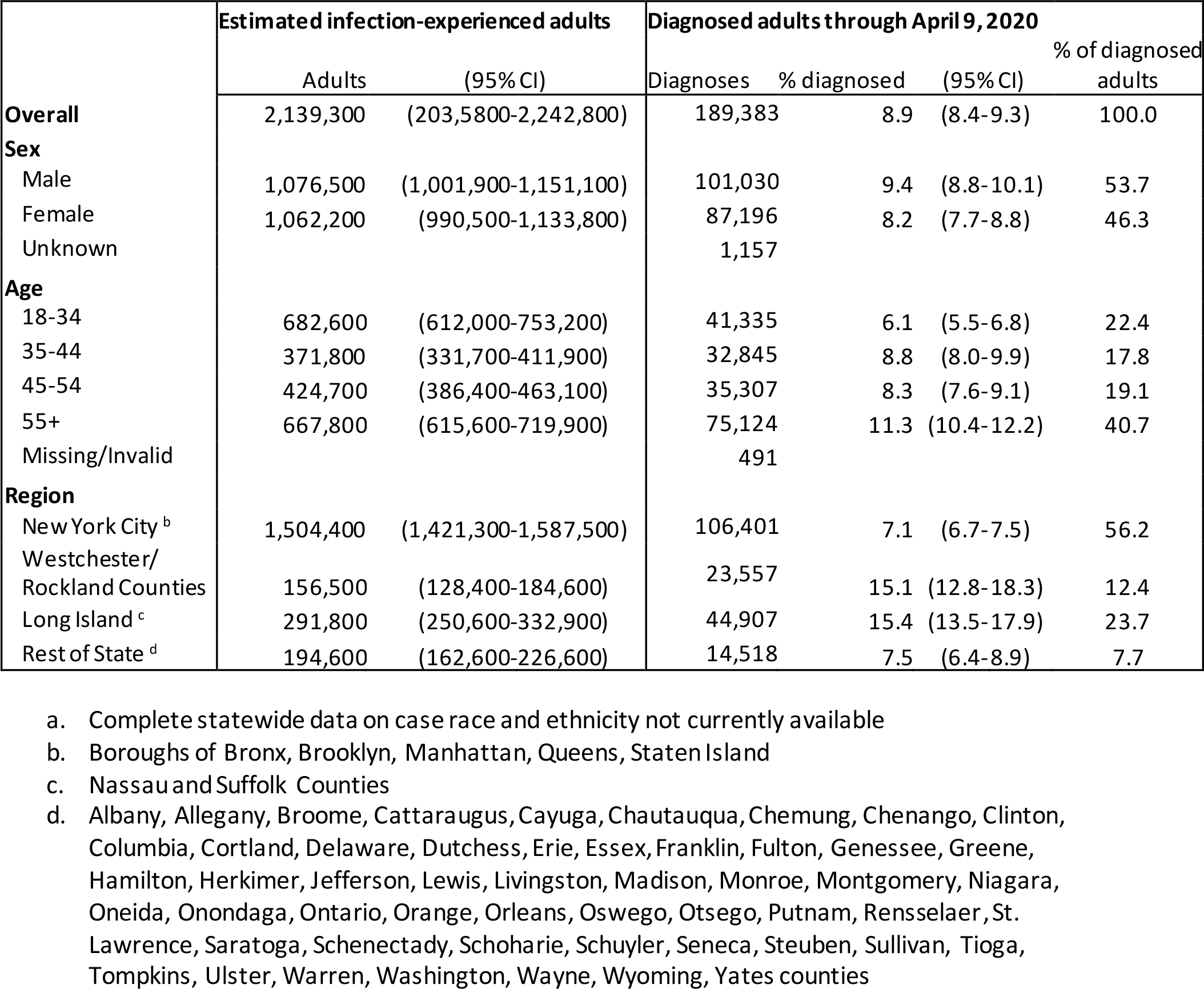
Estimated percentage of SARS-CoV-2 infections diagnosed^a^

## Discussion

From the largest US SARS-CoV-2 serosurvey to-date, we estimated that over 2 million adult NYS residents were infected through the end of March. Our findings estimate the extent of transmission of and community experience with SARS-CoV-2, particularly in the NYC metropolitan region. Despite large numbers of persons acquiring SARS-CoV-2, this represents only 14.0% of adult residents, suggesting that, even in this COVID-19 epicenter, the epidemic is substantially below the ~70% US herd immunity threshold.^22^ Against this remaining epidemic potential, ongoing vigilance through rigorous and extensive epidemic monitoring, testing, and contact tracing are necessary components for predicting, preventing, and/or mitigating a second epidemic wave, consistent with state and federal guidance for reopening.^5,23^ This vigilance is needed even in the rest of NYS outside the metropolitan region, portions of which are beginning the first phases of reopening in NYS, and where lowest cumulative incidence suggests the highest proportion susceptible.

Our finding of higher cumulative incidence in the regions of the NYC metropolitan area, particularly NYC, is consistent with the known distribution of diagnoses. Further, in these regions of high urbanicity, significant racial/ethnic disparities in infection history were found, with minority communities experiencing disproportionate risk. The drivers of greater COVID-19 risk and disparities in urban areas continue to be studied, but may relate to population density and the mechanisms by which transportation, employment, housing, and other socioeconomic or environmental factors shape opportunities for transmission.^24-26^ A recent NYS study on a random sample of COVID-19 hospitalizations showed limited racial/ethnic differences in clinical outcomes, suggesting that observed differences in mortality by race and ethnicity are likely driven by the different infection histories reported here.^3,27^ Research is needed to understand the drivers of increased COVID-19 risk experienced by minority communities, followed by actions to improve health equity.

The finding that over 8.9% of adults were diagnosed reveals both the opportunities for further expansion of diagnostic testing in NYS, yet in context of far higher diagnosis and testing levels than other US settings suggests substantial progress to-date.^1,11^ Compared to all persons with infection history, there was a higher representation of males and those over age 55 among diagnosed persons. Given the lower reactivity rates observed among this age group, our results expand observations from previous studies that older adults may be more likely to exhibit symptoms or illness or be more likely to seek care.^27-30^

Strengths of our study include a large sample, which contained 0.1% of the adult NYS population, and a systematic sampling approach in one of the only open public venues in the state, where a necessary commodity is purchased. Although a convenience sample, survey weights adjusted for biased demographic/geographic representation, and we further adjusted results for assay performance, under varied scenarios. Our study may nevertheless be limited by residual non-representativeness of the underlying population. This includes potential undersampling of persons from vulnerable groups who might be less likely to go grocery shopping. For this to impact our findings, those remaining home would need to have differential antibody prevalence compared to their age/sex/racial-ethnic/regional group peers. If persons staying at home had lower prevalence due to self-isolation, our study’s cumulative incidence would be a slight overestimate. Further, our sample did not include those who have died from COVID-19 or those who reside in long term care facilities which have been differentially impacted, causing a slight underestimate, nor those in the hospital or at-home due to COVID-19 illness, some of whom would be expected to have detectable antibodies.^31,32^ Such actively symptomatic persons would be expected to be a small portion of the cumulative infection burden since the outbreak’s commencement, and given most would have been infected after March 29, their exclusion also likely causes observed values to be overestimated.

Although data are limited on the potential for self-selection to alter our results, a recent Icelandic study found comparable prevalence when participants were tested following online self-registration vs. random invitation.^13^ This finding, in conjunction with our systematic community intercept approach, suggest that this bias may be small, outside of outright non-response. Although data were not captured, field workers reported very-high willingness to participate due to the public concern over COVID-19, supporting low non-response. Results presented may differ from publicly discussed preliminary estimates, given both our inclusion of more participants and analytic adjustments for test characteristics. Timeframes utilized for cumulative infections and diagnoses are approximate, being based on the evolving SARS-CoV-2 immunological and testing literature, with the 10-day sampling period during a linear-growth phase of the epidemic.

The findings of this study suggest extensive SARS-CoV-2 transmission in NYS and highlight the remaining opportunities for prevention and diagnosis. As the epidemic grows in other regions of the country, this study offers a potential model for other jurisdictions to monitor their epidemic. Estimates of cumulative incidence can be combined with diagnostic totals, or other epidemic markers such as mortality, to provide a holistic epidemic view during a time of unprecedented pandemic and to best craft high-impact approaches to prevention, containment, treatment and mitigation.

## Data Availability

Data are not publicly available

## Acknowledgements

The co-authors wish to acknowledge the following contributors to this work. Data preparation team members Janson Ganns, Peter Cichetti, Melissa Kamal, Alison Pingelski and Mary McCormick. Michelle Cummings for data management. Amy Kelly for literature review contributions. The New York State Antibody Sampling and Testing Team, including the Call Center team who placed thousands of phone calls to deliver test results and to collect missing demographic data. Office of Quality and Patient Safety team members James Kirkwood and Meng Wu. Eric Hall at Emory University for map assistance. We thank Dr. William Lee for assistance with assay development and validation, Jean Rock and the Wadsworth Center COVID-19 serology tea m especially Rachel Bievenue, Seth Blumerman, Theresa Hattenrath, Jim Long, Kate Mastraccio, Erica Miller, Katie Nemeth, and Alyssa Sossei, and numerous members of Wadsworth Center’s Newborn Screening Program especially Beth Vogel, Michele Caggana and Rhonda Hamel.

## Notes

### Competing Interest Statement

The authors have declared no competing interest.

### Funding Statement

No funding

### Author Declarations

New York State Department of Health IRB approved this study

